# Antibody Response after Second-dose of ChAdOx1-nCOV (Covishield™^®^) and BBV-152 (Covaxin™^®^) among Health Care Workers in India: Final Results of Cross-sectional Coronavirus Vaccine-induced Antibody Titre (COVAT) study

**DOI:** 10.1101/2021.06.02.21258242

**Authors:** Awadhesh Kumar Singh, Sanjeev Ratnakar Phatak, Ritu Singh, Kingshuk Bhattacharjee, Nagendra Kumar Singh, Arvind Gupta, Arvind Sharma

**Author notes:** **Corresponding Author:** A. K. Singh, G.D Hospital & Diabetes Institute, Kolkata - 700013, India; e mail. **Declarations of interest:** We wish to confirm that there are no known conflicts of interest associated with this publication and there has been no financial support for this work. **Authorship:** All authors meet the International Committee of Medical Journal Editors (ICMJE) criteria for authorship and take responsibility for the integrity of the work. They confirm that this paper will not be published elsewhere in the same form, in English or in any other language, including electronically.

## Abstract

**Background:** We assessed the humoral immune response after the completion of two doses of both ChAdOx1-nCOV (Covishield™) and BBV-152 (Covaxin™) vaccines in Indian health care workers (HCW).

**Methods:** A Pan-India, Cross-sectional, Coronavirus Vaccine-induced Antibody Titre (COVAT) study was conducted that measured SARS-CoV-2 anti-spike binding antibody quantitatively, 21 days or more after the first and second dose of two vaccines in both severe acute respiratory syndrome (SARS-CoV-2) naïve and recovered HCW. Primary aim was to analyze antibody response (seropositivity rate and median [inter-quartile range, IQR] antibody titre) following each dose of both vaccines and its correlation to age, sex, blood group, body mass index (BMI) and comorbidities. Here we report the final results of anti-spike antibody response after the two completed doses.

**Results:** Among the 515 HCW (305 Male, 210 Female), 95.0% showed seropositivity after two doses of both vaccines. Of the 425 Covishield and 90 Covaxin recipients, 98.1% and 80.0% respectively, showed seropositivity. However, both seropositivity rate and median (IQR) rise in anti-spike antibody was significantly higher in Covishield vs. Covaxin recipient (98.1 vs. 80.0%; 127.0 vs. 53 AU/mL; both p<0.001). This difference persisted in 457 SARS-CoV-2 naïve cohorts and propensity-matched (age, sex and BMI) analysis of 116 cohorts. While no difference was observed in relation to sex, BMI, blood group and any comorbidities; people with age >60 years or those with type 2 diabetes had a significantly lower seropositivity rates. Both vaccine recipients had similar solicited mild to moderate adverse events and none had severe or unsolicited side effects. In SARS-CoV-2 naïve cohorts, sex, presence of comorbidities, and vaccine type were independent predictors of antibody positivity rate in multiple logistic regression analysis.

**Conclusions:** Both vaccines elicited good immune response after two doses, although seropositivity rates and median anti-spike antibody titre was significantly higher in Covishield compared to Covaxin arm.

**Highlights:**

1. This study evaluated the humoral antibody response after 2 doses of SARS-CoV-2 vaccine Covishield™ and Covaxin™ in Indian health-care workers.
2. Combined results of both vaccines showed 95% seropositivity to anti-spike antibody, 21-36 days after the second completed dose.
3. Seropositivity rates were higher in Covishield recipients compared to Covaxin in the propensity-matched analysis of SARS-CoV-2 naïve cohorts.
4. Gender, presence of comorbidities and the type of vaccine received were independent predictors of antibody response after the second dose.

## 1. Introduction

Nation-wide vaccination against the Severe Acute Respiratory Syndrome Coronavirus 2 (SARS-CoV-2) infection, the cause of Coronavirus disease 2019 (COVID-19) pandemic is currently ongoing across the globe. Vaccination with two candidate vaccines namely Covishield™ and Covaxin™ in India has been started from January 16, 2021 after the Emergency Use Approval (EUA). Covishield™ (ChAdOx1-nCOV or AZD1222, acquired from Oxford University and AstraZeneca, manufactured by Serum Institute of India, Pune) is a recombinant replication-deficient chimpanzee adenovirus-vectored vaccine encoding SARS-CoV-2 spike antigen produced in genetically modified human embryonic kidney (HEL) 293 cells. Each dose (0.5 ml) of Covishield contains 5 x 10^10^ viral spike particles. Covaxin™ (BBV-152, manufactured by Bharat Biotech, Hyderabad in collaboration with Indian Council of Medical Research [ICMR], India) is a ß-propiolactone inactivated whole virion vaccine having all structural SARS-CoV-2 antigens adjuvanted by imidazoquinoline Toll-like receptor 7/8 (TLR 7/8) agonist to boost cell-mediated immunity. Each dose (0.5 ml) of Covaxin contains 6 µg dose of whole virion inactivated corona virus antigen strain NIV-2020-770. While, early data from available phase 3 randomized clinical trials (RCTs) suggested that these two vaccines are safe and effective [1-3], there is still a paucity of information as to how much and how long, these novel vaccines can elicit an immune response, both at humoral and cellular level in real-world settings. Moreover, the antibody kinetics after the completion of 2 doses of Covishield and Covaxin and thereafter, over a period of time is less well known. We have reported the preliminary results of ongoing Cross-sectional Coronavirus Vaccine-induced Antibody Titre (COVAT) study that assessed the anti-spike antibody humoral response 21-day after the first dose but before the second dose of both Covishield and Covaxin [4]. In this analysis, we report the binding anti-spike antibody kinetics after the completion of second dose of two vaccines from the ongoing COVAT study.

## 2. Methods

### 2.1 Study design and participants

This report followed the Strengthening the Reporting of Observational Studies in Epidemiology (STROBE) reporting guideline for cross-sectional studies [5]. COVAT study is an ongoing, pan-India, cross-sectional study that was approved by the ethical committee of Thakershy Charitable Trust, Ahmedabad, Gujarat, India. Written informed consent were taken from all the participants who participated in this study, voluntarily. All adult health care workers of more than 18 years of age who received the first dose of vaccine were eligible to participate in this study including those who had recovered from the COVID-19 in the recent past (> 6 weeks before the first dose). Individuals with current confirmed SARS-CoV-2 infection and those diagnosed within 6-weeks were excluded from the study. All subjects have received the two doses of either vaccine 0.5 ml intramuscularly in deltoid region of arm. Data collection for this analysis started since the January 16, 2021 (first day of vaccination amongst HCW) until May 15, 2021 (data-lock date). India had nearly 25 million cases of COVID-19 with an average case reported per day ranged from 0.1-0.4 million during this period, with a peak of >0.4 million case on a single day on May 7, 2021.

### 2.2 Measurements

Clinical data was collected from all eligible participants including age, sex, blood groups, body mass index (BMI), past history of confirmed SARS-CoV-2 infection, any comorbidities, presence of diabetes mellitus (type 1 [T1DM] and type 2 [T2DM]), hypertension (HTN) including its duration and treatment received, dyslipidemia, ischemic heart disease (IHD), chronic kidney disease (CKD) and cancer. Additional data was collected for any adverse events post-vaccination after second dose and subsequent SARS-CoV-2 infection (breakthrough infections). Accordingly, all participants were instructed to record and report the severity of conventional or solicited adverse events (fever, pain, induration, swelling, redness, muscle pain, headache, fatigue, rash, pruritus or acute allergic reaction) to site investigators occurring within a week of vaccination. Severity of solicited adverse events was graded as nil, mild (recovered within 24 to 48-hour) to moderate (>48-hour to <7-days) and severe (requiring hospitalization), depending upon the intensity and duration of adverse event(s). Similarly, a record of unexpected or unsolicited adverse events of bleeding, thrombo-embolic episode, bells’ palsy, seizure or other neurological manifestations, occurring from day 0 to 6 month is also being captured. Data collection for unsolicited adverse events and breakthrough infections is still ongoing and will be continued for 6-months after the second dose. Breakthrough infections are reported as per the given proforma that include-timing and severity, defined as mild (in-house or hospital treatment not requiring oxygen therapy), moderate (requiring oxygen supplementation but not requiring assisted ventilation, and severe (requiring ventilator).

As reported previously, anti-spike antibody titre is being measured at four time-points: day 21 after the first dose until the day before the second dose; day 21-28 of second dose, day 83-97 (3-months) and day 173-187 (6-months) after the second dose. Second blood samples (5 ml) were collected from eligible from day 21-36 after the second dose of each vaccine. An additional 7 days for second blood sample was allowed due to widespread lockdown. All samples were collected as either serum or plasma using EDTA vials from each participant and analyzed at Central laboratory of Neuberg, Supratech at Ahmedabad, Gujarat, India. The IgG antibodies to SARS- CoV-2 directed against the spike protein (S-antigen, both S1 and S2 protein) were assayed with LIASON^®^ S1/S2 quantitative antibody detection kit (DiaSorin Saluggia, Italy) using indirect chemiluminescence immunoassay (CLIA). The diagnostic sensitivity of this kit has been reported to be 97.4% (95% Confidence Interval [CI], 86.8-99.5%) >15 days after SARS-CoV-2 infection with a specificity of 98.5% (95% CI, 97.5-99.2%) as per manufacturer’s protocol. Antibody levels >15.0 arbitrary unit (AU)/mL were considered as sero-positive, while antibody level ≤15 AU/mL are considered as seronegative, as per manufacturer’s kit. This kit has found to have a concordance to neutralizing antibody (NAb) done by Plaque Reduction Neutralization Test (PRNT) with a positive agreement of 94.4% (95% CI, 88.8-97.2%) and negative agreement of 97.8% (95% CI, 94.4-99.1%) at a cut-off of >15 AU/mL. The lower and upper limit of this quantitative spike antibody kit is 3.8 and 400 AU/mL respectively, as per manufacture’s brochure [6].

### 2.3 Statistical analysis

Descriptive and inferential statistical analysis has been carried out for the present study. Normality of the data was assessed by Shapiro-Wilk test and visually by QQ plot for Covishield and Covaxin subgroups. Data on continuous scale was presented as Median (Interquartile range, IQR) and categorical data were presented as number (%). A two-sided P value of < 0.05 was considered as statistically significant. Chi-square test was used to find the significance of study parameters on categorical scale between two or more groups. Mann-Whitney test was utilized to assess two non-parametric groups and Kruskal-Wallis test was used to compare the differences among two or multiple data group for data on continuous scale. To compare the antibody kinetics between two vaccines, we also carried out a propensity-matched comparison of the two groups. A propensity score was generated taking into consideration age, sex and BMI of the SARS-CoV-2 naïve participants. Participants having similar scores were matched and two groups were compared accordingly. Moreover, multiple logistic regression analysis was also conducted to find out whether any independent factors were associated with a blunted response to vaccine in anti-spike antibody generation following first dose of vaccination. Additionally, in order to provide more reliable information on the independent predictors of antibody levels, explanatory variables were dummy coded and a log transformation of the outcome variable was done to perform multiple regression analysis. The significant predictors were back transformed by taking an antilog to facilitate information of the data. Entire statistical analysis was carried out with Statistical Package for Social Sciences (SPSS Complex Samples) Software Version 22.0 for windows, SPSS Inc., Chicago, IL, USA, with Microsoft Word and Excel being used to generate graphs and tables.

## 3. Results

**Figure 1** depicts the flow diagram of participants disposition that entered in to final analysis after the exclusion of 37 cohorts from the first preliminary analysis. Five hundred and fifteen participants who received second dose of either of the two vaccines, had complete set of data including anti-spike antibody. The mean age of the participants was 44.8 ± 13.1 years, with 59.2% males (305/515) and 40.8% females (210/515). Out of 515 participants, 437 (84.9%) were aged ≤ 60 years and 78 (15.1%) were of age > 60 years. While 24.3% (125/515) had one or more comorbidities, 10.1% had T2DM, 18.3% had HTN, 4.7% had dyslipidemia and 2.5% had IHD. We had no participant having T1DM, CKD or cancer in our study cohort. Among all participants, 11.3% (58/515) had past history of confirmed SARS-CoV-2 (2-week prior to the first dose of vaccine) who completed two doses of either of the vaccine. While there was no imbalance in baseline characteristics, a significantly larger proportion of patients had higher BMI, any comorbidities and past history of COVID-19 in Covishield compared to the Covaxin arm. **Table 1** summarizes the baseline characteristics of 515 cohorts.

**Table 1:** Baseline subject characteristics (N=515).

| Characteristics | Overall, N=515 | Covishield, N= 425 | Covaxin, N=90 | P value |
| --- | --- | --- | --- | --- |
| Age |  |  |  |  |
| Age (in years), Mean ± SD | 44.82 ± 13.09 | 45.15 ± 12.26 | 43.44 ± 16.04 | 0.286 |
| Age ≤ 60 years, n (%) | 437 (84.9) | 361 (84.9) | 76 (84.4) | 0.905 |
| Age > 60 years, n (%) | 78 (15.1) | 64 (15.1) | 14 (15.6) |  |
| Sex |  |  |  |  |
| Male, n (%) | 305 (59.2) | 252 (59.3) | 53 (58.9) | 0.943 |
| Female, n (%) | 210 (40.8) | 173 (40.7) | 37 (41.1) |  |
| Body-mass Index (BMI) |  |  |  |  |
| BMI (kg/m<sup>2</sup>), Mean ± SD | 25.42 ± 4.43 | 25.96 ± 4.87 | 24.02 ± 3.66 | 0.001 |
| BMI < 25 kg/m<sup>2</sup>, n (%) | 353 (68.5) | 287 (67.5) | 66 (73.3) | 0.531 |
| BMI 25-29.9 kg/m<sup>2</sup>, n (%) | 96 (18.6) | 81 (19.1) | 15 (16.7) |  |
| BMI ≥ 30 kg/m<sup>2</sup>, n (%) | 66 (12.8) | 57 (13.4) | 9 (10) | |
| Blood group |  |  |  |  |
| A+ve | 104 (20.2) | 86 (20.2) | 18 (20) | 0.319 |
| B+ve | 148 (28.7) | 132 (31.1) | 16 (17.8) |  |
| AB+ve | 29 (5.6) | 24 (5.6) | 5 (5.6) |  |
| O+ve | 205 (39.8) | 155 (36.5) | 50 (55.6) |  |
| A-ve | 7 (1.4) | 7 (1.6) |  |  |
| B-ve | 9 (1.7) | 9 (2.1) |  |  |
| AB-ve | 2 (0.4) | 2 (0.5) |  |  |
| O-ve | 11 (2.1) | 10 (2.4) | 1 (1.1) |  |
| Comorbidities |  |  |  |  |
| Any Co-morbidities, n (%) | 125 (24.3) | 114 (26.8) | 11 (12.2) | 0.003 |
| No Co-morbidities, n (%) | 390 (75.7) | 311 (73.2) | 79 (87.8) |  |
| Type 2 diabetes mellitus (T2DM) |  |  |  |  |
| T2DM, n (%) | 52 (10.1) | 46 (10.8) | 6 (6.7) | 0.234 |
| No T2DM, n (%) | 463 (89.9) | 379 (89.2) | 84 (93.3) |  |
| Duration of T2DM |  |  |  |  |
| T2DM Duration < 5 years, n (%) | 8 (1.6) | 8 (1.9) |  | 0.409 |
| T2DM Duration 5-10 years, n (%) | 30 (5.8) | 25 (5.9) | 5 (5.6) |  |
| T2DM Duration > 10 years, n (%) | 14 (2.7) | 13 (3.1) | 1 (1.1) |  |
| T2DM-Optimum control, n (%) | 51 (9.9) | 45 (10.6) | 6 (6.7) | 0.470 |
| T2DM-No optimum control, n (%) | 1 (0.1) | 1 (0.2) | 0 |  |
| *T2DM Management* |  |  |  |  |
| Monotherapy, n (%) | 15 (2.9) | 13 (3.1) | 2 (2.2) | 0.615 |
| Combination therapy, n (%) | 32 (6.2) | 29 (6.8) | 3 (3.3) |  |
| Insulin, n (%) | 1 (0.02) | 0 | 1 (1.1) |  |
| No medication, n (%) | 4 (0.1) | 4 (0.9) |  |  |
| *Hypertension (HTN)* |  |  |  |  |
| HTN, n (%) | 94 (18.3) | 75 (17.6) | 19 (21.1) | 0.440 |
| No HTN, n (%) | 421 (81.7) | 350 (82.4) | 71 (78.9) |  |
| *Duration of HTN* |  |  |  |  |
| HTN Duration < 5 years, n (%) | 27 (5.2) | 24 (5.6) | 3 (3.3) | 0.053 |
| HTN Duration 5-10 years, n (%) | 46 (8.9) | 31 (7.3) | 15 (16.7) |  |
| HTN Duration > 10 years, n (%) | 21 (4.1) | 20 (4.7) | 1 (1.1) |  |
| *HTN Management* |  |  |  |  |
| RAAS Blockers, n (%) | 40 (7.8) | 37 (8.7) | 3 (3.3) | 0.158 |
| CCBs, n (%) | 21 (4.1) | 19 (4.5) | 2 (2.2) |  |
| Combined, n (%) | 20 (3.9) | 18 (4.2) | 2 (2.2) |  |
| No Medicine, n (%) | 2 (0.4) | 1 (0.2) | 1 (1.1) |  |
| *Dyslipidemia* |  |  |  |  |
| Dyslipidemia, n (%) | 24 (4.7) | 24 (5.6) | 0 | 0.062 |
| No Dyslipidemia, n (%) | 491 (95.3) | 401 (94.4) | 90 (100) |  |
| *Ischemic heart disease (IHD)* |  |  |  |  |
| IHD, n (%) | 13 (2.5) | 13 (3.1) |  | 0.093 |
| No IHD, n (%) | 502 (97.5) | 412 (96.9) | 90 (100) |  |
| *History of SARS-CoV-2 infection* |  |  |  |  |
| Past H/o COVID-19, n (%) | 58 (11.3) | 55 (12.9) | 3 (3.3) | 0.009 |
| No Past H/o COVID-19, n (%) | 457 (88.7) | 370 (87.1) | 87 (96.7) |  |

p<0.05 considered as statistically significant, p computed by chi-square test or independent sample t-test to test the significance between Covaxin and Covishield

**Figure 1.**
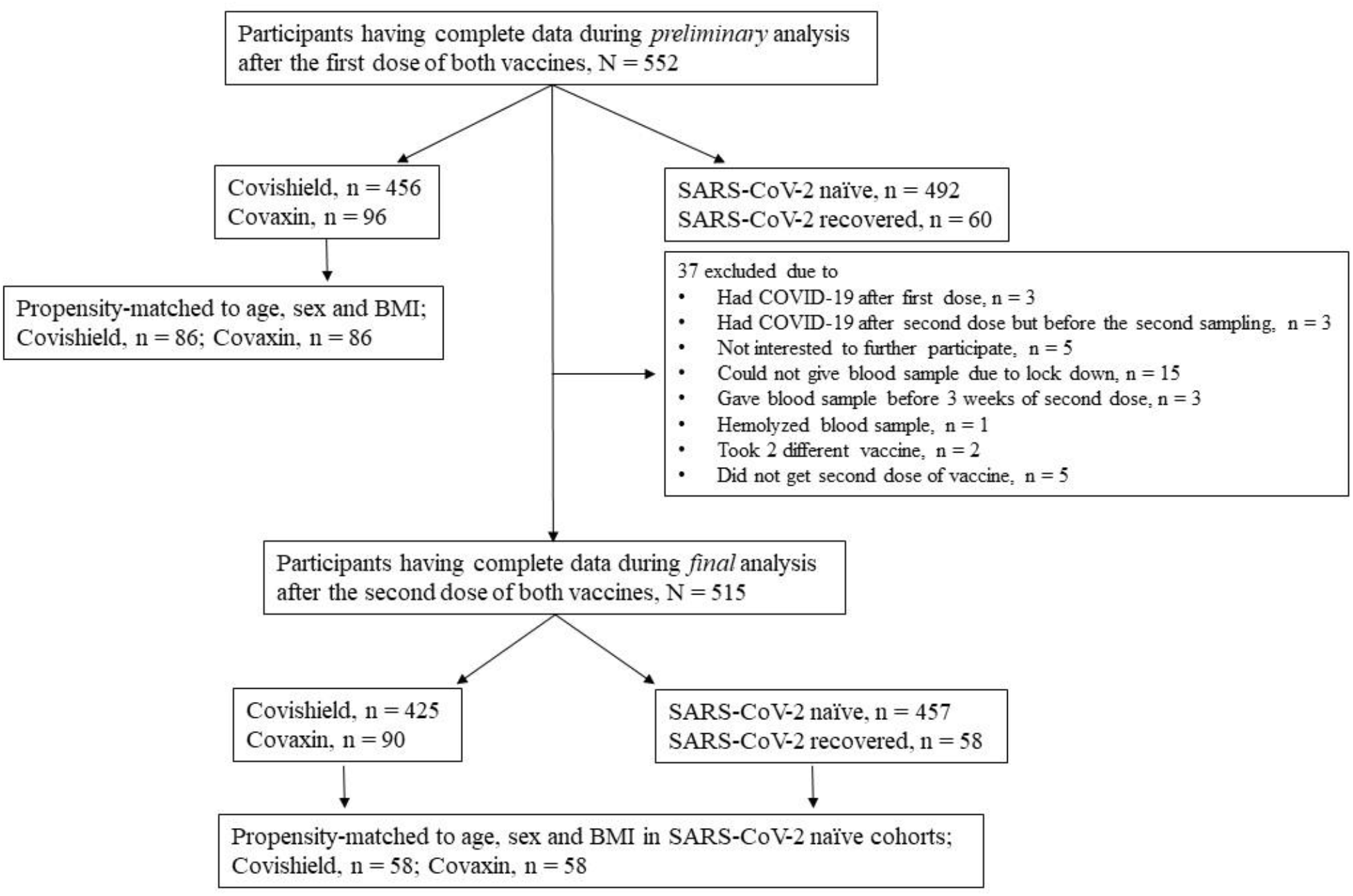
Flow diagram for participants disposition after first and second dose of each vaccine.

### 3.1 SARS-COV-2 spike antibody positivity rates after first dose of two vaccine

A total of 515 (305 male, 210 female) participants’ data was analyzed. Out of these 425 and 90 had received both doses of Covishield and Covaxin respectively. Overall, 95.0% (489/515) had seropositivity (defined as anti-spike antibody titre >15AU/mL measured at day 21-36 after the second dose) for anti-spike antibody. Notably, the seropositivity rate was significantly higher in Covishield vs. Covaxin recipients (98.1 vs. 80.0% respectively, p<0.001). Seropositivity rate was significantly (p<0.001) more in participants with age ≤60 years (96.3%) vs. >60 years (87.2%), in overall cohorts, primarily contributed by Covishield recipients. While no difference in seropositivity rate was observed for gender, BMI and presence of any comorbidities in overall cohorts between the two arms, females compared to males (100.0 vs. 96.8%, p=0.02), people with BMI <25 compared to >25 Kg/m2 (99.3 vs. 95.7%, p=0.03) and the presence of any comorbidities compared to those without (95.6 vs. 99.0%, p=0.02), had a significantly higher seropositivity rate in Covishield arm. Amongst the captured co-morbidities, people with T2DM had a significantly lower seropositivity rate compared to those without (84.6 vs. 96.1%, p=0.002) in overall cohort, and this was consistent in both Covishield (91.3 vs. 98.9%, p<0.001) and Covaxin recipients (63.6 vs. 82.3%, p=0.003), respectively. Moreover, people with T2DM of shorter duration (<5 years) had a higher seropositivity rate (100.0 vs. 89.5%, p<0.001) compared to those having longer duration (>5 year), and this was consistently observed in both Covishield and Covaxin recipients. Although no difference of seropositivity rate was observed in people with HTN, those having shorter (<5 years) duration had a significantly higher seropositivity (100.0 vs. 88.0%, p=0.008) vs. longer duration (>5 years). Intriguingly, no significant difference in seropositivity rate was observed between SARS-CoV-2 naïve vs. people with past history of SARS-CoV-2 infection (94.3 vs. 100.0%, p=0.06) after the two complete doses of both vaccines. No differential antibody seropositivity rate was observed in relation to types of blood group, presence or absence of dyslipidemia, IHD and treatment regime of DM and HTN (**Table 2**).

**Table 2:** Seropositivity to anti-spike antibody after (day 21-36) the second dose of either vaccine

| Characteristics | Overall, N= 515 |  | Covishield, N= 425 |  | Covaxin, N= 90 |  |
| --- | --- | --- | --- | --- | --- | --- |
|  | Seropositivity Rate | P | Seropositive Rate | P | Seropositivity Rate | P |
| *Age* |  |  |  |  |  |  |
| Age ≤ 60 years, n (%) | 421/437 (96.3) | 0.001 | 358/361 (99.2) | <0.001 | 63/76 (82.9) | 0.110 |
| Age > 60 years, n (%) | 68/78 (87.2) |  | 59/64 (92.2) |  | 9/14 (64.3) |  |
| *Sex* |  |  |  |  |  |  |
| Male, n (%) | 286/305 (93.8) | 0.140 | 244/252 (96.8) | 0.018 | 42/53 (79.2) | 0.830 |
| Female, n (%) | 203/210 (96.7) |  | 173/173 (100) |  | 30/37 (81.1) |  |
| *Body-mass Index (BMI)* |  |  |  |  |  |  |
| BMI < 25 kg/m<sup>2</sup> | 338/353 (95.8) | 0.441 | 285/287 (99.3) | 0.028 | 53/66 (80.3) | 0.984 |
| BMI 25-29.9 kg/m<sup>2</sup> | 90/96 (93.8) |  | 78/81 (96.3) |  | 12/15 (80) |  |
| BMI ≥ 30 kg/m<sup>2</sup> | 61/66 (92.4) |  | 54/57 (94.7) |  | 7/9 (77.8) |  |
| *Blood group* |  |  |  |  |  |  |
| A+ve, n (%) | 99/104 (95.2) | 0.799 | 86/86 (100) | 0.569 | 13/18 (72.2) | 0.396 |
| B+ve, n (%) | 142/148 (81.8) |  | 127/132 (96.2) |  | 15/16 (93.8) |  |
| AB+ve, n (%) | 26/29 (89.7) |  | 23/24 (95.8) |  | 3/5 (60) |  |
| O+ve, n (%) | 193/205 (94.1) |  | 153/155 (98.7) |  | 40/50 (80) |  |
| A-ve, n (%) | 7/7 (100) |  | 7/7 (100) |  |  |  |
| B-ve, n (%) | 9/9 (100) |  | 9/9 (100) |  |  |  |
| AB-ve, n (%) | 2/2 (100) |  | 2/2 (100) |  |  |  |
| O-ve, n (%) | 11/11 (100) |  | 10/10 (100) |  | 0/1 (0) |  |
| *Comorbidities* |  |  |  |  |  |  |
| Any Co-morbidities, n (%) | 116/125 (92.8) | 0.207 | 109/114 (95.6) | 0.021 | 7/11 (63.6) | 0.148 |
| No Co-morbidities, n (%) | 373/390 (95.6) |  | 308/311 (99) |  | 65/79 (82.3) |  |
| *Type 2 diabetes mellitus (T2DM)* |  |  |  |  |  |  |
| T2DM, n (%) | 44/52 (84.6) | 0.002 | 42/46 (91.3) | <0.001 | 2/6 (33.3) | 0.003 |
| No T2DM, n (%) | 445/463 (96.1) |  | 375/379 (98.9) |  | 70/84 (83.3) |  |
| *Duration of T2DM* |  |  |  |  |  |  |
| T2DM Duration < 5 years, n (%) | 8/8 (100) | <0.001 | 8/8 (100) | <0.001 | 0 | 0.008 |
| T2DM Duration 5-10 years, n (%) | 23/30 (76.7) |  | 21/25 (84) |  | 2/5 (40) |  |
| T2DM Duration > 10 years, n (%) | 13/14 (92.9) |  | 13/13 (100) |  | 0/1 (0) |  |
| *Control of T2DM* |  |  |  |  |  |  |
| T2DM-Optimum control, n (%) | 43/51 (84.3) | 0.369 | 41/45 (91.1) | 0.568 | 2/6 (33.3) | 0.762 |
| T2DM-No optimum control, n (%) | 1/1 (100) |  | 1/1 (100) |  | 0 (0/0) |  |
| *T2DM Management* |  |  |  |  |  |  |
| Monotherapy, n (%) | 14/15 (93.3) | 0.151 | 13/13 (100) | 0.201 | 1/2 (50) | 0.103 |
| Combination therapy, n (%) | 26/32 (81.2) |  | 26/29 (89.7) |  | 0/3 (0) |  |
| Insulin, n (%) | 0 |  | 0 |  | 0 (0) |  |
| No medication, n (%) | 4/5 (80) |  | 3/4 (75) |  | 1/1 (100) |  |
| *Hypertension (HTN)* |  |  |  |  |  |  |
| HTN, n (%) | 86/94 (91.5) | 0.090 | 72/75 (96) | 0.137 | 14/19 (73.7) | 0.438 |
| No HTN, n (%) | 403/421 (95.7) |  | 345/350 (98.6) |  | 58/71 (81.7) |  |
| *Duration of HTN* |  |  |  |  |  |  |
| HTN Duration < 5 years, n (%) | 27/27 (100) | 0.008 | 24/24 (100) | 0.010 | 3/3 (100) | 0.152 |
| HTN Duration 5-10 years, n (%) | 39/46 (84.8) |  | 28/31 (90.3) |  | 11/15 (73.3) |  |
| HTN Duration > 10 years, n (%) | 20/21 (95.2) |  | 20/20 (100) |  | 0/1 (0) |  |
| *HTN Management* |  |  |  |  |  |  |
| RAAS Blockers, n (%) | 37/40 (92.5) | 0.958 | 35/37 (94.6) | 0.352 | 2/3 (66.7) | 0.695 |
| CCBs, n (%) | 20/21 (95.2) |  | 18/19 (94.7) |  | 2/2 (100) |  |
| Combined, n (%) | 19/20 (95) |  | 18/18 (100) |  | 1/2 (50) |  |
| No Medicine, n (%) | 2/2 (100) |  | 1/1 (100) |  | 1/1 (100) |  |
| *Dyslipidemia* |  |  |  |  |  |  |
| Dyslipidemia, n (%) | 23/24 (95.8) | 0.954 | 23/24 (95.8) | 0.692 | 0 | * |
| No Dyslipidemia, n (%) | 466/491 (94.9) |  | 394/401 (98.2) |  | 72/90 (80) |  |
| *Ischemic heart disease (IHD)* |  |  |  |  |  |  |
| IHD, n (%) | 13/13 (100) | 0.400 | 13/13 (100) | 0.612 | 0 | * |
| No IHD, n (%) | 476/502 (94.8) |  | 404/412 (98.1) |  | 72/90 (80) |  |
| *History of SARS-CoV-2 infection* |  |  |  |  |  |  |
| Past H/o COVID-19, n (%) | 58/58 (100) | 0.062 | 55/55 (100) | 0.271 | 3/3 (100) | 0.378 |
| No Past H/o COVID-19, n (%) | 431/457 (94.3) |  | 362/370 (97.8) |  | 69/87 (79.3) |  |
| *Types of vaccines* |  |  |  |  |  |  |
| Covishield, n (%) | 417/425 (98.1) | <0.001 |  |  |  |  |
| Covaxin, n (%) | 72/90 (80) |  |  |  |  |  |
| p<0.05 considered as statistically significant, p computed by chi-square test, BMI- body mass index, T2DM- type 2 diabetes mellitus, HTN- hypertension, RAAS- Renin angiotensin aldosterone system, CCB- calcium channel blocker, IHD- ischemic heart disease, COVID-19- coronavirus disease 2019, * - No statistics is computed |  |  |  |  |  |  |

Since past history of SARS-CoV-2 infection can interfere with seropositivity to anti-spike antibody, we additionally analyzed the seropositivity rate after excluding SARS-CoV-2 recovered (n=58) cohorts. Importantly, in this analysis of 457 cohorts’ results were similar and consistent. Seropositivity rate after the two complete doses was significantly higher in Covishield compared to Covaxin recipients (97.8 vs. 79.3%; p <0.001) (**Supplementary table 1**).

### 3.2 Comparison of SARS-CoV-2 spike antibody between Covishield and Covaxin after propensity matched analysis

In the propensity-matched analysis of 116 SARS-CoV-2 naïve cohorts (58 participants in each arm) after the adjustment for age, sex and BMI; seropositivity rates were significantly higher in Covishield arm compared to the Covaxin (98.3% vs. 82.8%, p=0.004). Age ≤60 years had a higher seropositivity rate vs. >60-years (98.5 vs. 79.6%, p=0.001) and presence of T2DM was associated with a lesser seropositivity rate compared to those without (92.0 vs. 50.0%, p=0.005) (**Table 3**).

**Table 3:**
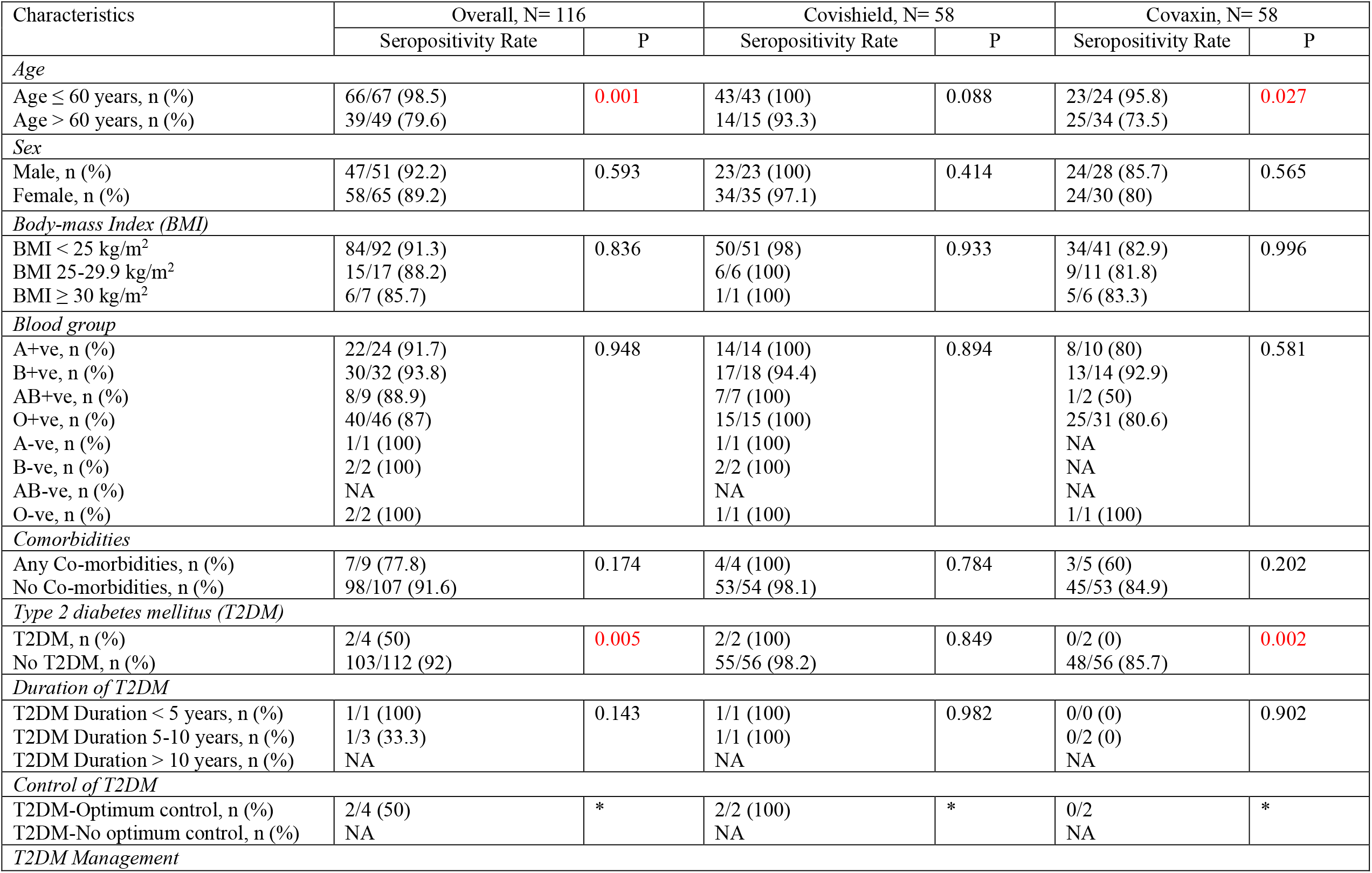
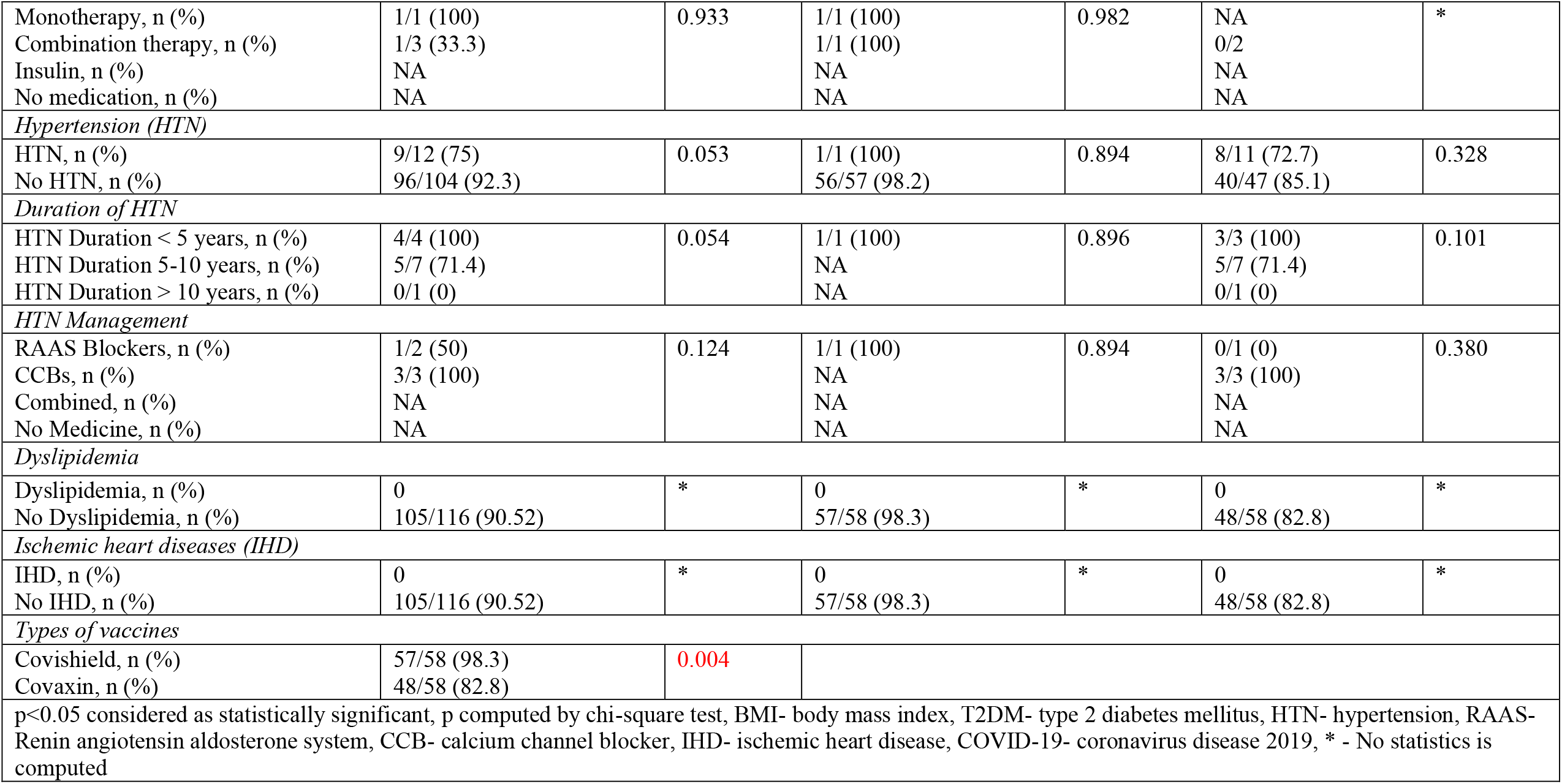
Seropositivity to anti-spike antibody after (day 21-36) the second dose of either vaccine in propensity score (age, sex, BMI) matched analysis of SARS-CoV-2 naïve cohorts

| Characteristics | Overall, N= 116 |  | Covishield, N= 58 |  | Covaxin, N= 58 |  |
| --- | --- | --- | --- | --- | --- | --- |
|  | Seropositivity Rate | P | Seropositivity Rate | P | Seropositivity Rate | P |
| *Age* |  |  |  |  |  |  |
| Age ≤ 60 years, n (%) | 66/67 (98.5) | 0.001 | 43/43 (100) | 0.088 | 23/24 (95.8) | 0.027 |
| Age > 60 years, n (%) | 39/49 (79.6) |  | 14/15 (93.3) |  | 25/34 (73.5) |  |
| *Sex* |  |  |  |  |  |  |
| Male, n (%) | 47/51 (92.2) | 0.593 | 23/23 (100) | 0.414 | 24/28 (85.7) | 0.565 |
| Female, n (%) | 58/65 (89.2) |  | 34/35 (97.1) |  | 24/30 (80) |  |
| *Body-mass Index (BMI)* |  |  |  |  |  |  |
| BMI < 25 kg/m<sup>2</sup> | 84/92 (91.3) | 0.836 | 50/51 (98) | 0.933 | 34/41 (82.9) | 0.996 |
| BMI 25-29.9 kg/m<sup>2</sup> | 15/17 (88.2) |  | 6/6 (100) |  | 9/11 (81.8) |  |
| BMI ≥ 30 kg/m<sup>2</sup> | 6/7 (85.7) |  | 1/1 (100) |  | 5/6 (83.3) |  |
| *Blood group* |  |  |  |  |  |  |
| A+ve, n (%) | 22/24 (91.7) | 0.948 | 14/14 (100) | 0.894 | 8/10 (80) | 0.581 |
| B+ve, n (%) | 30/32 (93.8) |  | 17/18 (94.4) |  | 13/14 (92.9) |  |
| AB+ve, n (%) | 8/9 (88.9) |  | 7/7 (100) |  | 1/2 (50) |  |
| O+ve, n (%) | 40/46 (87) |  | 15/15 (100) |  | 25/31 (80.6) |  |
| A-ve, n (%) | 1/1 (100) |  | 1/1 (100) |  | NA |  |
| B-ve, n (%) | 2/2 (100) |  | 2/2 (100) |  | NA |  |
| AB-ve, n (%) | NA |  | NA |  | NA |  |
| O-ve, n (%) | 2/2 (100) |  | 1/1 (100) |  | 1/1 (100) |  |
| *Comorbidities* |  |  |  |  |  |  |
| Any Co-morbidities, n (%) | 7/9 (77.8) | 0.174 | 4/4 (100) | 0.784 | 3/5 (60) | 0.202 |
| No Co-morbidities, n (%) | 98/107 (91.6) |  | 53/54 (98.1) |  | 45/53 (84.9) |  |
| *Type 2 diabetes mellitus (T2DM)* |  |  |  |  |  |  |
| T2DM, n (%) | 2/4 (50) | 0.005 | 2/2 (100) | 0.849 | 0/2 (0) | 0.002 |
| No T2DM, n (%) | 103/112 (92) |  | 55/56 (98.2) |  | 48/56 (85.7) |  |
| *Duration of T2DM* |  |  |  |  |  |  |
| T2DM Duration < 5 years, n (%) | 1/1 (100) | 0.143 | 1/1 (100) | 0.982 | 0/0 (0) | 0.902 |
| T2DM Duration 5-10 years, n (%) | 1/3 (33.3) |  | 1/1 (100) |  | 0/2 (0) |  |
| T2DM Duration > 10 years, n (%) | NA |  | NA |  | NA |  |
| *Control of T2DM* |  |  |  |  |  |  |
| T2DM-Optimum control, n (%) | 2/4 (50) | * | 2/2 (100) | * | 0/2 | * |
| T2DM-No optimum control, n (%) | NA |  | NA |  | NA |  |
| *T2DM Management* |  |  |  |  |  |  |
| Monotherapy, n (%) | 1/1 (100) | 0.933 | 1/1 (100) | 0.982 | NA | * |
| Combination therapy, n (%) | 1/3 (33.3) |  | 1/1 (100) |  | 0/2 |  |
| Insulin, n (%) | NA |  | NA |  | NA |  |
| No medication, n (%) | NA |  | NA |  | NA |  |
| *Hypertension (HTN)* |  |  |  |  |  |  |
| HTN, n (%) | 9/12 (75) | 0.053 | 1/1 (100) | 0.894 | 8/11 (72.7) | 0.328 |
| No HTN, n (%) | 96/104 (92.3) |  | 56/57 (98.2) |  | 40/47 (85.1) |  |
| *Duration of HTN* |  |  |  |  |  |  |
| HTN Duration < 5 years, n (%) | 4/4 (100) | 0.054 | 1/1 (100) | 0.896 | 3/3 (100) | 0.101 |
| HTN Duration 5-10 years, n (%) | 5/7 (71.4) |  | NA |  | 5/7 (71.4) |  |
| HTN Duration > 10 years, n (%) | 0/1 (0) |  | NA |  | 0/1 (0) |  |
| *HTN Management* |  |  |  |  |  |  |
| RAAS Blockers, n (%) | 1/2 (50) | 0.124 | 1/1 (100) | 0.894 | 0/1 (0) | 0.380 |
| CCBs, n (%) | 3/3 (100) |  | NA |  | 3/3 (100) |  |
| Combined, n (%) | NA |  | NA |  | NA |  |
| No Medicine, n (%) | NA |  | NA |  | NA |  |
| *Dyslipidemia* |  |  |  |  |  |  |
| Dyslipidemia, n (%) | 0 | * | 0 | * | 0 | * |
| No Dyslipidemia, n (%) | 105/116 (90.52) |  | 57/58 (98.3) |  | 48/58 (82.8) |  |
| *Ischemic heart diseases (IHD)* |  |  |  |  |  |  |
| IHD, n (%) | 0 | * | 0 | * | 0 | * |
| No IHD, n (%) | 105/116 (90.52) |  | 57/58 (98.3) |  | 48/58 (82.8) |  |
| *Types of vaccines* |  |  |  |  |  |  |
| Covishield, n (%) | 57/58 (98.3) | 0.004 |  |  |  |  |
| Covaxin, n (%) | 48/58 (82.8) |  |  |  |  |  |
| p<0.05 considered as statistically significant, p computed by chi-square test, BMI- body mass index, T2DM- type 2 diabetes mellitus, HTN- hypertension, RAAS- Renin angiotensin aldosterone system, CCB- calcium channel blocker, IHD- ischemic heart disease, COVID-19- coronavirus disease 2019, * - No statistics is computed |  |  |  |  |  |  |

### 3.3 Assessment of SARS-CoV-2 spike antibody quantity after two complete doses of Covishield and Covaxin

The median (IQR) rise in anti-spike antibody was significantly higher (p<0.001) in Covishield recipients 127.0 (70.5-268.5) AU/mL, compared to Covaxin arm 53.0 (22.3-131.0) AU/mL. Significantly higher median values of anti-spike antibody were observed in people having age ≤60 vs. >60 years (120.0 vs. 92.0 AU/mL, p=0.03), primarily skewed by higher value in Covishield arm (131.0 vs. 113.0 AU/mL respectively, p=0.03). In the overall cohort, the females had a significantly higher median antibody titre compared to the males (131.0 vs. 112.0 AU/mL, p=0.01), primarily driven by Covishield arm. Although presence of any comorbidities was not associated with any differential antibody titre, presence of HTN was associated with a significantly attenuated median antibody titre compared to those without (95.0 vs. 124.0 AU/mL, p=0.002), particularly in the Covishield arm. Indeed, past history of SARS-CoV-2 infection elicited a significantly greater median antibody titre, compared to SARS-CoV-2 naïve cohorts (400.0 vs. 110.0 AU/mL, p <0.001), irrespective of the type of vaccine received. **Table 4** summarizes the results of antibody titre across all groups. Box and whisker plot in **Figure 2** depicts the log antibody titre for different study parameters that were significantly different after the two complete doses of vaccines, while **Supplementary figure 1** complements the absolute antibody (median, IQR) titre in same subgroups.

**Table 4:**
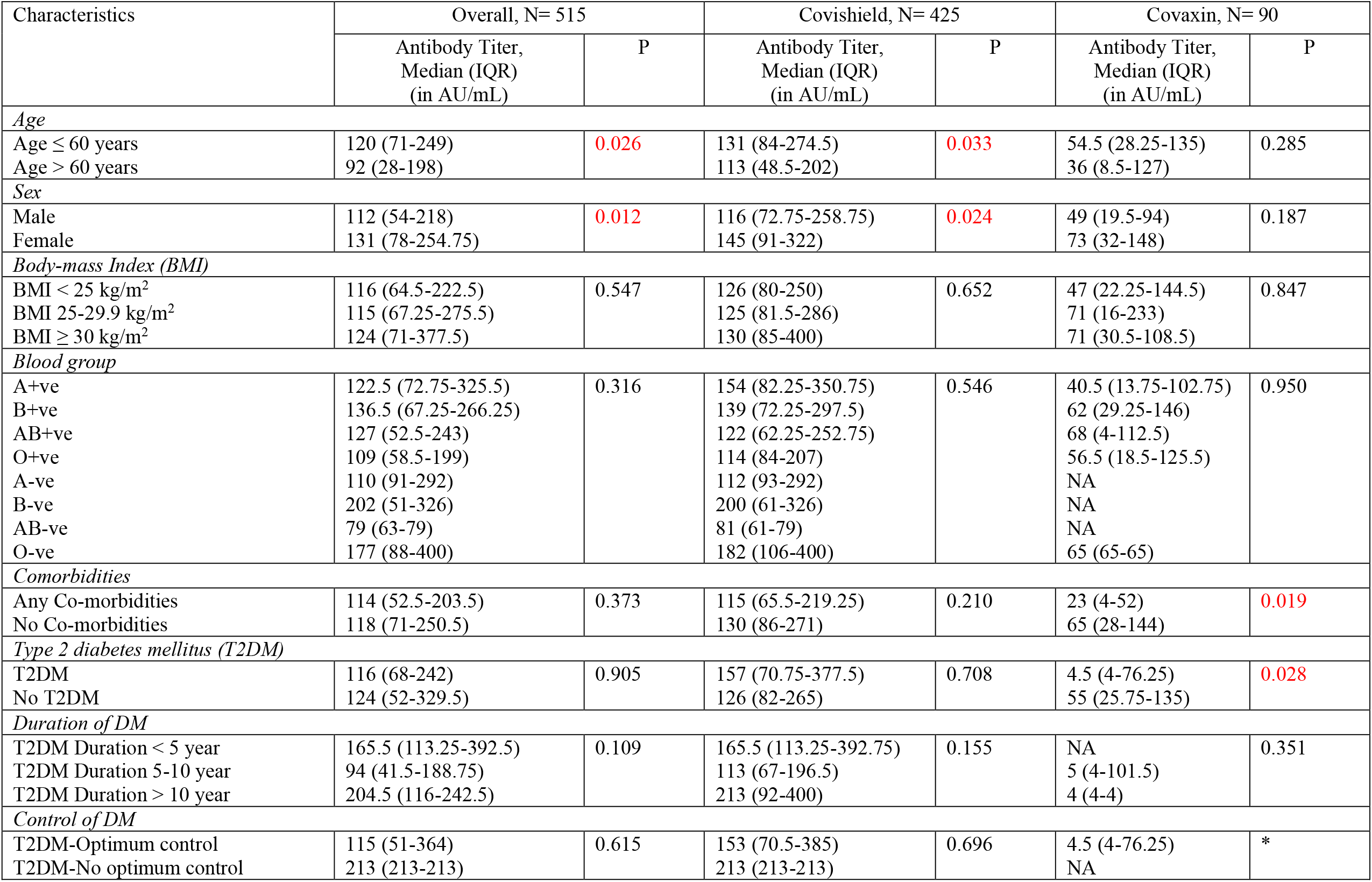
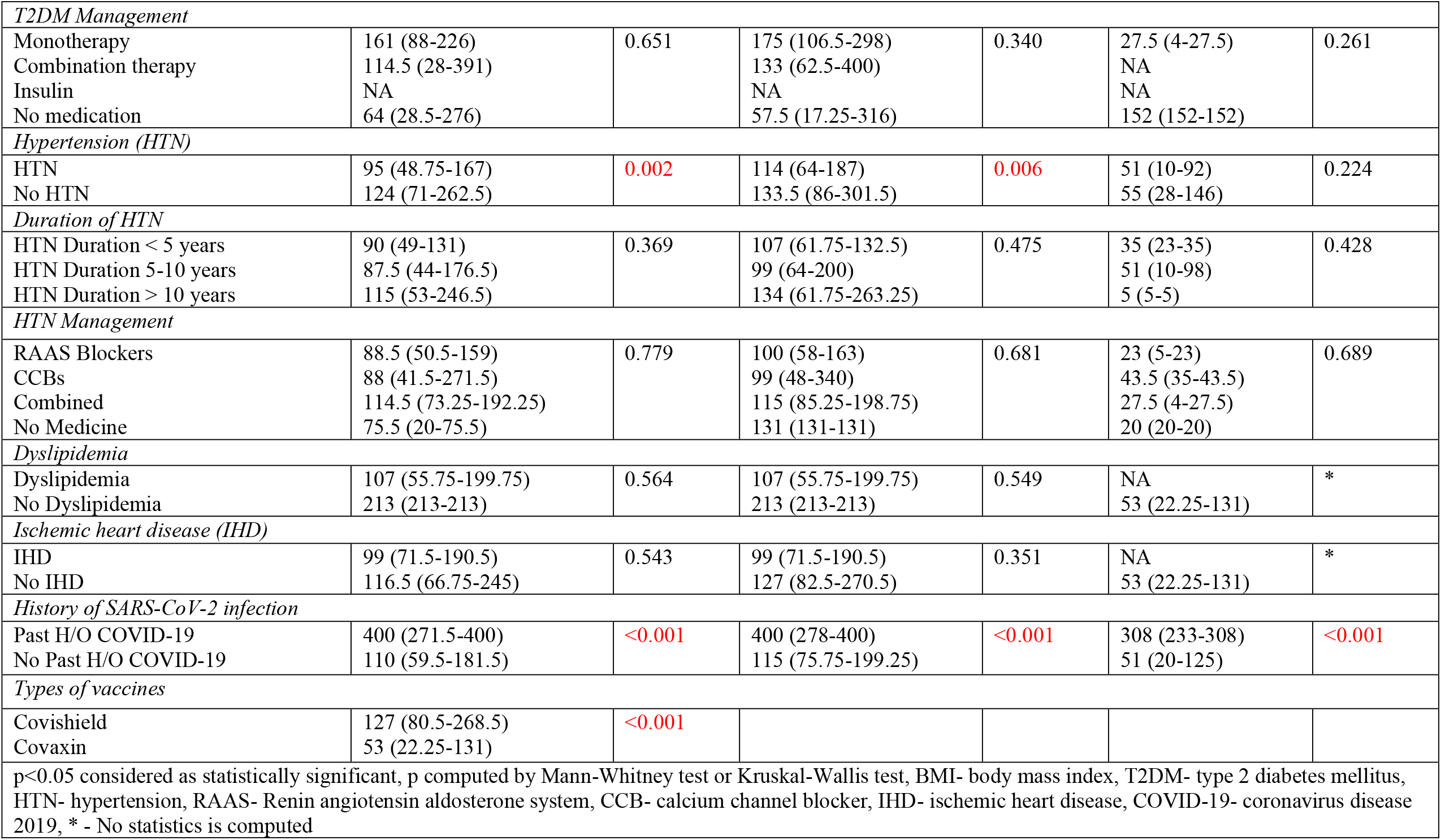
Median (interquartile range) anti-spike antibody titre after (day 21-36) the second dose of either vaccine

| Characteristics | Overall, N= 515 |  | Covishield, N= 425 |  | Covaxin, N= 90 |  |
| --- | --- | --- | --- | --- | --- | --- |
|  | Antibody Titer,<br>Median (IQR)<br>(in AU/mL) | P | Antibody Titer,<br>Median (IQR)<br>(in AU/mL) | P | Antibody Titer,<br>Median (IQR)<br>(in AU/mL) | P |
| *Age* |  |  |  |  |  |  |
| Age ≤ 60 years | 120 (71-249) | 0.026 | 131 (84-274.5) | 0.033 | 54.5 (28.25-135) | 0.285 |
| Age > 60 years | 92 (28-198) |  | 113 (48.5-202) |  | 36 (8.5-127) |  |
| *Sex* |  |  |  |  |  |  |
| Male | 112 (54-218) | 0.012 | 116 (72.75-258.75) | 0.024 | 49 (19.5-94) | 0.187 |
| Female | 131 (78-254.75) |  | 145 (91-322) |  | 73 (32-148) |  |
| *Body-mass Index (BMI)* |  |  |  |  |  |  |
| BMI < 25 kg/m<sup>2</sup> | 116 (64.5-222.5) | 0.547 | 126 (80-250) | 0.652 | 47 (22.25-144.5) | 0.847 |
| BMI 25-29.9 kg/m<sup>2</sup> | 115 (67.25-275.5) |  | 125 (81.5-286) |  | 71 (16-233) |  |
| BMI ≥ 30 kg/m<sup>2</sup> | 124 (71-377.5) |  | 130 (85-400) |  | 71 (30.5-108.5) |  |
| *Blood group* |  |  |  |  |  |  |
| A+ve | 122.5 (72.75-325.5) | 0.316 | 154 (82.25-350.75) | 0.546 | 40.5 (13.75-102.75) | 0.950 |
| B+ve | 136.5 (67.25-266.25) |  | 139 (72.25-297.5) |  | 62 (29.25-146) |  |
| AB+ve | 127 (52.5-243) |  | 122 (62.25-252.75) |  | 68 (4-112.5) |  |
| O+ve | 109 (58.5-199) |  | 114 (84-207) |  | 56.5 (18.5-125.5) |  |
| A-ve | 110 (91-292) |  | 112 (93-292) |  | NA |  |
| B-ve | 202 (51-326) |  | 200 (61-326) |  | NA |  |
| AB-ve | 79 (63-79) |  | 81 (61-79) |  | NA |  |
| O-ve | 177 (88-400) |  | 182 (106-400) |  | 65 (65-65) |  |
| *Comorbidities* |  |  |  |  |  |  |
| Any Co-morbidities | 114 (52.5-203.5) | 0.373 | 115 (65.5-219.25) | 0.210 | 23 (4-52) | 0.019 |
| No Co-morbidities | 118 (71-250.5) |  | 130 (86-271) |  | 65 (28-144) |  |
| *Type 2 diabetes mellitus (T2DM)* |  |  |  |  |  |  |
| T2DM | 116 (68-242) | 0.905 | 157 (70.75-377.5) | 0.708 | 4.5 (4-76.25) | 0.028 |
| No T2DM | 124 (52-329.5) |  | 126 (82-265) |  | 55 (25.75-135) |  |
| *Duration of DM* |  |  |  |  |  |  |
| T2DM Duration < 5 year | 165.5 (113.25-392.5) | 0.109 | 165.5 (113.25-392.75) | 0.155 | NA | 0.351 |
| T2DM Duration 5-10 year | 94 (41.5-188.75) |  | 113 (67-196.5) |  | 5 (4-101.5) |  |
| T2DM Duration > 10 year | 204.5 (116-242.5) |  | 213 (92-400) |  | 4 (4-4) |  |
| *Control of DM* |  |  |  |  |  |  |
| T2DM-Optimum control | 115 (51-364) | 0.615 | 153 (70.5-385) | 0.696 | 4.5 (4-76.25) | * |
| T2DM-No optimum control | 213 (213-213) |  | 213 (213-213) |  | NA |  |
| *T2DM Management* |  |  |  |  |  |  |
| Monotherapy | 161 (88-226) | 0.651 | 175 (106.5-298) | 0.340 | 27.5 (4-27.5) | 0.261 |
| Combination therapy | 114.5 (28-391) |  | 133 (62.5-400) |  | NA |  |
| Insulin | NA |  | NA |  | NA |  |
| No medication | 64 (28.5-276) |  | 57.5 (17.25-316) |  | 152 (152-152) |  |
| *Hypertension (HTN)* |  |  |  |  |  |  |
| HTN | 95 (48.75-167) | 0.002 | 114 (64-187) | 0.006 | 51 (10-92) | 0.224 |
| No HTN | 124 (71-262.5) |  | 133.5 (86-301.5) |  | 55 (28-146) |  |
| *Duration of HTN* |  |  |  |  |  |  |
| HTN Duration < 5 years | 90 (49-131) | 0.369 | 107 (61.75-132.5) | 0.475 | 35 (23-35) | 0.428 |
| HTN Duration 5-10 years | 87.5 (44-176.5) |  | 99 (64-200) |  | 51 (10-98) |  |
| HTN Duration > 10 years | 115 (53-246.5) |  | 134 (61.75-263.25) |  | 5 (5-5) |  |
| *HTN Management* |  |  |  |  |  |  |
| RAAS Blockers | 88.5 (50.5-159) | 0.779 | 100 (58-163) | 0.681 | 23 (5-23) | 0.689 |
| CCBs | 88 (41.5-271.5) |  | 99 (48-340) |  | 43.5 (35-43.5) |  |
| Combined | 114.5 (73.25-192.25) |  | 115 (85.25-198.75) |  | 27.5 (4-27.5) |  |
| No Medicine | 75.5 (20-75.5) |  | 131 (131-131) |  | 20 (20-20) |  |
| *Dyslipidemia* |  |  |  |  |  |  |
| Dyslipidemia | 107 (55.75-199.75) | 0.564 | 107 (55.75-199.75) | 0.549 | NA | * |
| No Dyslipidemia | 213 (213-213) |  | 213 (213-213) |  | 53 (22.25-131) |  |
| *Ischemic heart disease (IHD)* |  |  |  |  |  |  |
| IHD | 99 (71.5-190.5) | 0.543 | 99 (71.5-190.5) | 0.351 | NA | * |
| No IHD | 116.5 (66.75-245) |  | 127 (82.5-270.5) |  | 53 (22.25-131) |  |
| *History of SARS-CoV-2 infection* |  |  |  |  |  |  |
| Past H/O COVID-19 | 400 (271.5-400) | <0.001 | 400 (278-400) | <0.001 | 308 (233-308) | <0.001 |
| No Past H/O COVID-19 | 110 (59.5-181.5) |  | 115 (75.75-199.25) |  | 51 (20-125) |  |
| *Types of vaccines* |  |  |  |  |  |  |
| Covishield | 127 (80.5-268.5) | <0.001 |  |  |  |  |
| Covaxin | 53 (22.25-131) |  |  |  |  |  |
| p<0.05 considered as statistically significant, p computed by Mann-Whitney test or Kruskal-Wallis test, BMI- body mass index, T2DM- type 2 diabetes mellitus, HTN- hypertension, RAAS- Renin angiotensin aldosterone system, CCB- calcium channel blocker, IHD- ischemic heart disease, COVID-19- coronavirus disease 2019, * - No statistics is computed |  |  |  |  |  |  |

**Figure 2.**
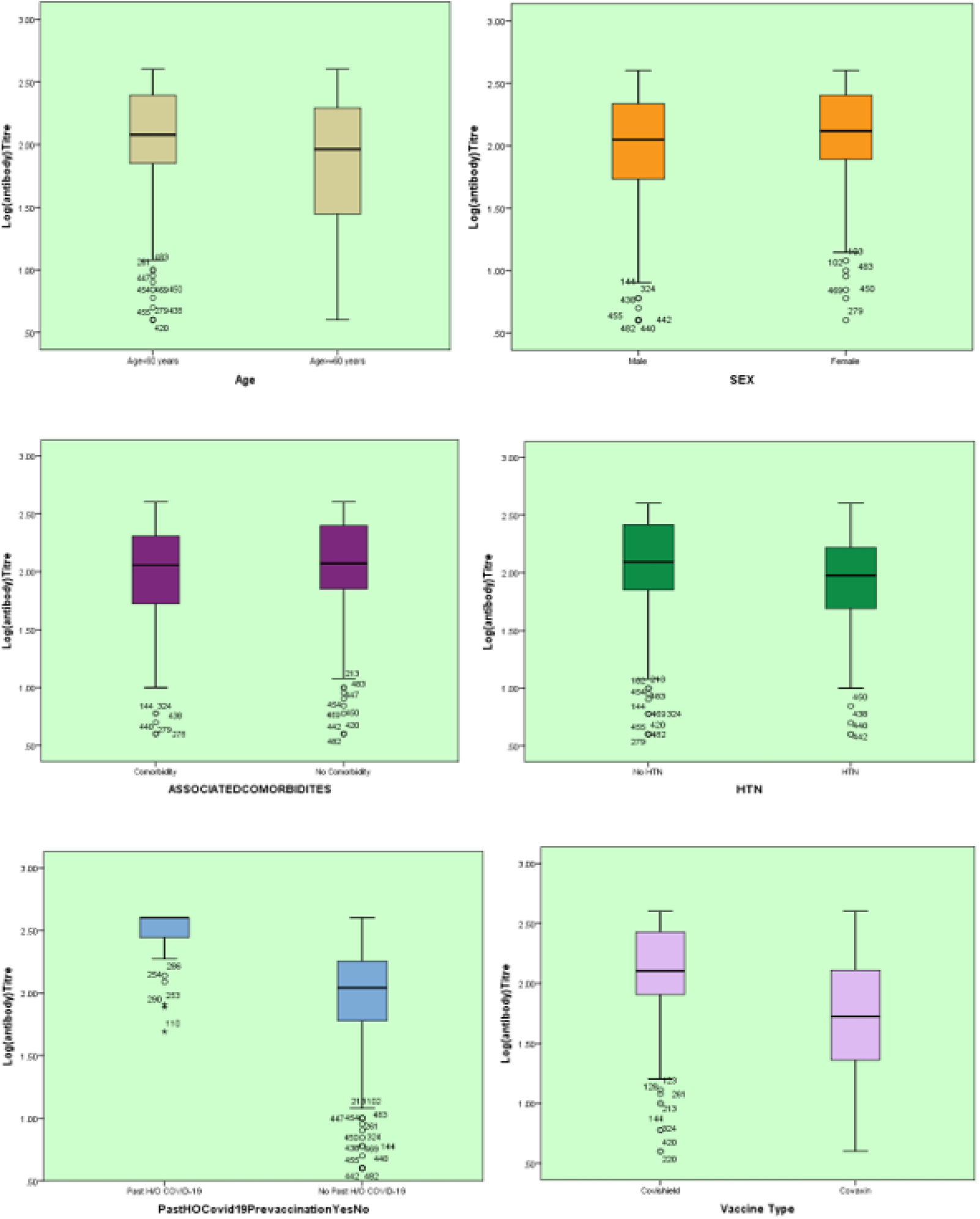
Box and whisker plot demonstrating log antibody titer by study parameters.

Since past history of COVID-19 can influence the antibody titre level, in a repeat analysis of 457 cohorts after excluding cohorts who had past history of COVID-19 (n=58), results were similar and consistent. The median (IQR) rise in anti-spike antibody titre was significantly higher in Covishield recipients compared to Covaxin (115.5 vs. 51.0 AU/mL, p <0.001), 21-36 days after the completion of second dose in SARS-CoV-2 naïve participants (**Supplementary table 2**).

### 3.4 Independent variables associated with responder vs. non-responders to spike antibody

Multiple logistic regression analysis (to identify the independent predictors for non-responder rate based on development of SARS-CoV-2 Anti-spike antibody) suggests that amongst all the variables analyzed, three variables such as-sex, presence of comorbidities, and vaccine type were independent predictors of antibody response rate. Associated co-morbidities were significantly related to an increase in the non - responder rate by more than 4 folds (Odds Ratio [OR] 4.68; 95% CI, 1.382 - 15.839; p=0.013), male gender also significantly increased the non-responder rate by more than 3 folds (OR 3.37; 95% CI, 1.221 - 9.321; p=0.019). Notably, recipients of Covishield had a significantly lower non-responder rate by 41% (OR 0.59; 95% CI, 0.021 - 0.659; p<0.001), compared to those who received Covaxin (**Table 5**).

**Table 5:** Multiple logistic regression to identify the independent predictors for non-seropositivity rate after second dose (based on development of SARS-CoV-2 Anti-spike antibody).

| Variables in the Final Equation |  |  |  |  |  |  |  |  |
| --- | --- | --- | --- | --- | --- | --- | --- | --- |
|  | B | S.E. | Wald | df | P value | Exp(B) | 95% C.I. for EXP(B) |  |
|  |  |  |  |  |  |  | Lower | Upper |
| Age | -.425 | .472 | .810 | 1 | .368 | .654 | .259 | 1.650 |
| Sex (ref= female) | 1.216 | .518 | 5.503 | 1 | .019 | 3.374 | 1.221 | 9.321 |
| BMI | -.205 | .280 | .536 | 1 | .464 | .815 | .471 | 1.410 |
| Associated Comorbidities | 1.543 | .622 | 6.152 | 1 | .013 | 4.679 | 1.382 | 15.838 |
| Type 2 diabetes | -.552 | .620 | .793 | 1 | .373 | .576 | .171 | 1.941 |
| Hypertension | .718 | .595 | 1.458 | 1 | .227 | 2.051 | .639 | 6.579 |
| Ischemic heart disease | -.286 | .950 | .062 | 1 | .893 | .789 | .423 | 6.084 |
| Dyslipidaemia | .000 | .000 | 3.401 | 1 | .065 | 1.000 | 1.000 | 1.000 |
| Past H/O Covid-19 | .732 | .609 | 1.444 | 1 | .230 | 2.078 | .630 | 6.854 |
| Vaccine type (ref=Covaxin) | -2.825 | .522 | 29.296 | 1 | <0.001 | .590 | .021 | .659 |
| a. Variable(s) entered on regression step: Age, Sex, BMI (body mass index), Associated Comorbidities, Type 2 diabetes, Hypertension, Ischemic heart disease, Dyslipidaemia, Past H/O Covid-19, Covishield/Covaxin. |  |  |  |  |  |  |  |  |

In a multiple linear regression model with log transformed antibody levels as outcome variable, we found presence of co-morbidity, sex, vaccine type and past history of COVID-19 infection as independent predictors of antibody titre. The R value of the model represents the simple correlation and is 0.493, which indicates a moderate degree of correlation. The overall R^2^ value is 0.247 indicating 24.7% of the total variation in the dependent variable (antibody titre) can be explained by our model. The multiple regression analysis (to identify the independent predictors for SARS-CoV-2 Anti-spike antibody titre) suggests that amongst all the variables analyzed, four variables such as-presence of comorbidity, sex, past history of SARS-CoV-2 infection and vaccine type were independent predictors of antibody titre levels. While presence of comorbidity resulted in decrease in the antibody titre by 13% (β=0.87; 95% CI, 0.79-0.97; p=0.010), female gender was found to have 9% greater antibody titre compared to males (β=1.09; 95% CI:1.02-1.17; p=0.018). SARS-CoV-2 naïve participants were associated with significantly lower antibody titre by 37% (β=0.63; 95% CI, 0.57-0.71; p<0.001). Notably, recipients of Covaxin had a significantly lower antibody titre by 32% ((β=0.68; 95%CI, 0.62-0.74; p<0.001), as compared to those who received Covishield (**Supplementary table 3**).

### 3.5 Post-vaccination (second dose) adverse events

In the overall post-vaccination cohort, the Covishield recipients reported a higher incidence of mild to moderate side-effects in 18.1% (77/425) patients compared to 11.1% (10/90) patients in Covaxin arm after the second dose of vaccine, however, it didn’t reach statistical significance (p=0.11). Notably, in the propensity-matched cohort, any side-effects (mild to moderate) post vaccination was also similar (p=0.13) in the Covishield arm 8/58 (13.79%) vs. Covaxin arm 6/58(10.34%). No serious solicited and any unsolicited side effects were noted.

### 3.6 Post-vaccination SARS-CoV-2 infection

From Jan 16, 2021 (Day 1 of vaccination) until May 15, 2021 (Day 21-36 after second dose, data-locking date), a total of 30 (30/492, 6.1%) HCW have reported to have COVID-19 among SARS-CoV-2 naïve cohorts (n=492) after the first dose of vaccination. Of the 30 participants 4 had suspected (symptomatic and positive HRCT signs but negative to either RT-PCR or RAT) and 26 had confirmed (RT-PCR or RAT positive) COVID-19. Three had SARS-CoV-2 infection before the second dose, while 27 had acquired SARS-CoV-2 infection after the second dose. Breakthrough infections (defined as SARS-CoV-2 infection >2 weeks after the second dose) were reported in 4.9% (24/492) of cases following both vaccines. Breakthrough infections were noted in 5.5% (22/399) cohorts in Covishield and 2.2% (2/93) of Covaxin recipients. Majority had mild (28/30) to moderate (2/30) COVID-19 infections and all recovered. None of the cohort who had COVID-19 following either vaccine had severe COVID requiring mechanical ventilation (**Supplementary Table 4**).

## 4. Discussion

Summarily, this cross-sectional COVAT study reported an overall 95.0% (489/515) seropositivity rate after the two complete doses of both vaccines in entire cohorts that include both SARS-CoV-2 naïve and recovered individuals (Covishield 98.1% and Covaxin 80.0%, respectively). While seropositivity rates after two complete doses was 97.8% and 79.3% with Covishield and Covaxin, respectively in SARS - CoV-2 naïve individuals; 100% of cohorts with a past history of SARS-CoV-2 were seropositive after the two doses of both vaccines. Notably, while both vaccines showed an increase in seropositivity and median (IQR) anti-spike antibody titre after the second dose, Covaxin gained a significant increase in both seropositivity and antibody titre only after the two completed doses. Contrarily, Covishield showed a good seropositivity rate and a 4-fold rise in median antibody titre even after a single dose. One dose of either vaccine yielded a very high seropositivity and anti-spike antibody titre in SARS-CoV-2 recovered individuals (**Table 6**). There was no significant difference in seropositivity rate with regard to age, sex, BMI, blood group and any comorbidities including its duration and treatment, except that the participants with T2DM and T2DM or HTN of >5-year duration had a significantly less seropositive rate compared to those without and of <5-year duration, respectively. Importantly, median titre of antibody was significantly higher in females compared to the males. Notably, no difference in seropositivity rate was observed amongst SARS-CoV-2 naïve vs. SARS-CoV-2 recovered cohorts after the two completed doses of both vaccines, however, median (IQR) anti-spike antibody titre was significantly higher in later compared to the former. Any adverse side effects post-vaccination was similar in Covishield and Covaxin recipient and were mild to moderate in nature. Surprisingly, the seropositivity (responder) rate and median anti-spike antibody titre was significantly higher in Covishield recipients, compared to the Covaxin. Whether this differential finding between two vaccines is related to a lesser number of participants in Covaxin arm compared to the Covishield, or due to the difference in characteristics of participants, or due to the difference between the type of vaccine-vector-based vs. inactivated whole virion, or related to differential immunogenic response due to the difference in the loading dose of antigen in each vaccine - is not exactly known- and need further studies. Nevertheless, even in age-, sex- and BMI-matched propensity analysis, seropositivity rate and median anti-spike antibody titre was significantly higher with Covishield compared to Covaxin in SARS-CoV-2 naïve recipients. Notably, sex, presence of comorbidities, and type of vaccine used were an independent predictor of antibody response in multiple regression analysis.

**Table 6:** Comparison of seropositivity rate, median anti-spike antibody titre (IQR) after the first (21-days or more but before second dose) and second dose (day 21-36) in total, SARS-CoV-2 naïve, and recovered participants

| Characteristics | Total cohorts |  |  | SARS-CoV-2 naïve participant <sup>#</sup> |  |  | Past history of SARS-CoV-2 infection |  |  |
| --- | --- | --- | --- | --- | --- | --- | --- | --- | --- |
|  | Covishield, n/N | Covaxin, n/N | Odds ratio, [95% CI], P value | Covishield, n/N | Covaxin, n/N | Odds ratio, [95% CI], P value | Covishield, n/N | Covaxin, n/N | Odds ratio, [95% CI], P value |
| *First dose* | *N = 552* |  |  | *N = 492* |  |  | *N = 60* |  |  |
| Seropositivity rate (%) | 396/456, (86.8) | 42/96, (43.8) | 8.48 [5.22-13.79] * | 341/399, (85.5) | 39/93, (41.9) | 8.14 [4.95-13.38] * | 55/57 (96.5) | 3/3 (100) | 3.17 [0.13-79.60] ** |
| Median [IQR] anti-spike antibody titre (AU/mL) | 61.5 [30.0-119.5] | 6.0 [4.0-99.7] | <0.001 | 55.0 [28.0-94.0] | 5.0 [4.0-85.5] | <0.001 | 400.0 [298.0-400.0] | 260.0 [214.0-260.0] | 0.42 |
| *Second dose* | *N = 515* |  |  | *N = 457* |  |  | *N = 58* |  |  |
| Seropositivity rate (%) | 417/425, (98.1) | 72/90, (80.0) | 13.03 [5.46-31.09] * | 362/370, (97.8) | 69/87, (79.3) | 11.80 [4.94-28.22] * | 55/55 (100) | 3/3 (100) | \$ |
| Median [IQR] anti-spike antibody titre (AU/mL) | 127.0 [80.5-268.5] | 53.0 [22.3-131.0] | <0.001 | 115.0 [75.8-199.3] | 20.0 [51.0-125.0] | <0.001 | 400.0 [278.0-400.0] | 308.0 [233.0-308.0] | 0.64 |
| <sup>#</sup> May include asymptomatic and undiagnosed COVID-19<br><sup>\$</sup> No statistics computed as seropositivity rate is constant<br><sup>*</sup> P < 0.001<br><sup>**</sup> P = 0.48 | | | | | | | | | |

Our findings are similar to published evidence in randomized controlled trials (RCTs) and real-world studies, although only few studies correlated it with other variables such as age, sex, BMI or presence or absence of comorbidities. In phase 2 RCT, 96.6% (95% CI, 92.8-98.8) had seropositivity (defined as post-vaccination IgG anti-spike antibody titre 4-fold higher than the baseline) to IgG anti-spike antibody titre at day 56 (28-days after the second dose), measured by ELISA in 177 participants who received 6 mcg dose of Algel-IDMG Covaxin. Moreover, seroconversion rate to NAb measured by PRNT_50_ rose to 98.3% 28-days after the second dose, compared to 47.5% 28-days after the first dose. However, no difference in NAb titre was observed in relation to gender and age [7, 8]. In phase 1/2 RCT, two doses of ChAdOx1 nCoV-19 (Covishield) vaccine elicited a significant increase in anti-spike IgG antibody at day 56 (Median 639 EU, IQR 360-792) compared to its median titre (210.7 EU, 149.4-206.8) at 28-days, as measured by ELISA in 10 prime-boost participants [9]. Similarly, in phase 2/3 RCT there was a significant increase in anti-spike IgG antibody in 1593 cohorts who took 2 standard doses of ChAdOx1 nCoV-19 measured by multiplex immunoassay [1, 2]. In a cross-sectional community survey from England (REal-time Assessment of Community Transmission-2 program, REACT-2) that studied IgG anti-spike antibody kinetics after two doses of Pfizer-BioNTech vaccine, involving 971 participants, found 91.1% seropositivity across all age groups. There was also a consistent decreasing trend in IgG positivity with the increasing age in REACT-2 study [10]. A high IgG seropositivity of 90.1% to a single dose of Pfizer-BioNTech vaccine was observed in people with past confirmed or suspected COVID across all age groups. Notably, none of these studies reported the antibody response in relation to comorbidities. Recent studies have reported a significantly diminished anti-spike antibody titre in hemodialysis patients compared to healthy control even after the two completed doses of both Pfizer-BioNTech vaccine and Moderna vaccines [11-13]. One study found highly attenuated humoral response to anti-spike antibody after two completed doses of Pfizer-BioNTech vaccine in people receiving disease-modifying drugs such as Ocrelizumab and Fingolimod for the treatment of multiple sclerosis [14].

The larger question is whether humoral antibody response to a vaccine correlates with the efficacy (reduction in severity and mortality due to COVID-19). Although the correlation of antibody titre to the vaccine efficacy is less well understood, NAb targeting different epitopes of spike glycoprotein have been found to protect from COVID-19 with ChAdOx1 nCoV-19 (Covishield) and Moderna mRNA-1273 vaccine [19, 20]. Moreover, NAb titre following vaccination was highly correlated with the NAb titre in convalescent post-SARS-CoV-2 infection. Furthermore, Anti-spike antibody titre is reported to highly correlate with in-vitro virus neutralization test measured by PNT [15]. Indeed, a strong correlation (*r* range 0·87–0·94) between NAb responses measured by PNT against the spike glycoprotein and those detected by ELISAs have been reported in patients with RT-PCR confirmed COVID-19 [16]. Contrarily, a longitudinal study found no relationship between post-vaccination serum binding-antibody in SARS-CoV-2 naïve individuals [17]. Collectively, it is not exactly known as to what level of binding and neutralizing antibody protects human from COVID-19 [18].

To the best of our knowledge, this Pan-India cross-sectional COVAT study would be the first of its kind that has involved HCW from 13 States and 22 cities and reporting anti-spike antibody kinetics after two completed doses of two different vaccines. However, we also acknowledge several limitations. Firstly, in the present study, we have used a convenience sampling amounting to selection bias. A community-based study in a larger population with multi-stage sampling would be an ideal sampling method. Secondly, we used a binary logistic regression (to identify the predictors of non-response to vaccines) which primarily assumes linearity between the explanatory variable and the outcome variable, hence this model may miss out any predictor variable which may have non-linear relationship with the outcome variable. Thirdly, we have measured only anti-spike binding antibody and could not assess NAb and cell-mediated immune response such as Th-1 and Th-2 dependent antibody or cytokines (primarily due to the lack of standardized commercial labs in India). Fourth, we could not measure the baseline anti-spike antibody titre prior to the vaccination, because of logistic issue due to lockdown. Finally, two value of short-term anti-spike antibody as evaluated in this report may not necessarily predict the efficacy of vaccine, nor the absence of seropositivity confer failure of vaccine in absence of NAb and T-cell response assessment.

In conclusion, this cross-sectional study after the completion of two doses of both vaccines suggests that both vaccines induce seropositivity to anti-spike antigen in 95% of SARS-COV-2 naïve and recovered individuals after 3-weeks. Whether any real difference in inducing immunogenicity exists between two vaccines can only be meaningfully demonstrated through a head-to-head RCT.

## Data Availability

All the authors are responsible for the originality of this study. Original data can be shared from first author, if necessary, after a reasonable request.

## Acknowledgments

We would like to thank all the participants who volunteered for this study. We express our sincere gratitude and acknowledgment to our Indian regional coordinators for the smooth conduct of this study that include (in alphabetical order) – Drs. Akash Kumar Singh (Vadodara), Amit Gupta (Noida), Anuj Maheshwari (Lucknow), Arvind Kumar Ojha (Kolkata), Bhavtharini (Erode), B. Harish Kumar (Mysore), J K Sharma (New Delhi), Jayant Panda (Cuttack), Kavyachand Yalamudi (Guntur), Kiran Shah (Vadodara), M Gowri Sankar (Coimbatore), Manohar KN (Bangalore), Meena Chhabra (New Delhi), Pratap Jethwani (Rajkot), M Shunmugavelu (Trichy), Rajiv Kovil (Mumbai), Sunil Gupta (Nagpur), Subhash Kumar (Patna), Somnath (Hyderabad), Urman Dhruv (Ahmedabad). Our heartfelt thanks to Ms. Roma Dave (Dietician) and Dr. Priya Phatak (Ahmedabad) for keeping entire data up-to-date and confidential at every step. Special thanks to Dr. Bhavini Shah, Dr. Sandip Shah, and Dr. Krutarth Shah from Neuberg Supratech Laboratory, Ahmedabad, for generously supporting our cause.

## Contribution of authors

AKS and SRP conceptualized and designed the study. NKS, AG and AS monitored the study and captured the data at all point of time. AKS, KB and RS conducted the statistical analysis. AKS and RS wrote the first draft. AKS, KB and RS revised the manuscript. All authors gave their intellectual inputs while preparing the manuscript and agreed mutually to submit to this journal.

