## Supplemental Table 1-4 for "Antibody Response after Second-dose of ChAdOx1-nCOV (Covishield™^®^) and BBV-152 (Covaxin™^®^) among Health Care Workers in India: Final Results of Cross-sectional Coronavirus Vaccine-induced Antibody Titre (COVAT) study"

**Supplementary Table 1: Seropositivity rate to anti-spike antibody after the second dose of either vaccine in SARS-CoV-2 naïve cohorts**

| Characteristics | Overall, N= 457 | | Covishield, N= 370 | | Covaxin, N= 87 | |
| --- | --- | --- | --- | --- | --- | --- |
|  | Seropositivity Rate | P | Seropositivity Rate | P | Seropositivity Rate | P |
| *Age* | | | | | | |
| Age ≤ 60 years, n (%)  Age > 60 years, n (%) | 304/388 (78.4)  47/69 (68.1) | 0.063 | 276/315 (87.6)  40/55 (72.7) | 0.004 | 28/73 (38.4)  7/14 (50) | 0.416 |
| *Sex* | | | | | | |
| Male, n (%)  Female, n (%) | 200/270 (74.1)  151/187 (80.7) | 0.096 | 181/219 (82.6)  135/151 (89.4) | 0.070 | 19/51 (37.3)  16/36 (44.4) | 0.501 |
| *Body-mass Index (BMI)* | | | | | | |
| BMI < 25 kg/m^2^  BMI 25-29.9 kg/m^2^  BMI ≥ 30 kg/m^2^ | 249/320 (77.8)  62/83 (74.7)  40/54 (74.1) | 0.735 | 226/256 (88.3)  54/69 (78.3)  36/45 (80) | 0.061 | 23/64 (35.9)  8/14 (57.1)  4/9 (44.4) | 0.329 |
| *Blood group* | | | | | | |
| A+ve, n (%)  B+ve, n (%)  AB+ve, n (%)  O+ve, n (%)  A-ve, n (%)  B-ve, n (%)  AB-ve, n (%)  O-ve, n (%) | 67/88 (76.1)  104/131 (79.4)  17/24 (70.8)  141/188 (75)  6/6 (100)  6/8 (75)  2/2 (100)  8/10 (80) | 0.800 | 64/72 (88.9)  97/115 (84.3)  15/19 (78.9)  118/139 (84.9)  6/6 (100)  6/8 (75)  2/2 (100)  8/9 (88.9) | 0.822 | 3/16 (18.8)  7/16 (43.8)  2/5 (40)  23/49 (46.9)  0 (0)  0 (0)  0 (0)  0/1 (0) | 0.315 |
| *Comorbidities* | | | | | | |
| Any Co-morbidities, n (%)  No Co-morbidities, n (%) | 71/106 (67)  280/351 (79.8) | 0.006 | 70/95 (73.7)  246/275 (89.5) | <0.001 | 1/11 (9.1)  34/76 (44.7) | 0.022 |
| *Type 2 diabetes mellitus (T2DM)* | | | | | | |
| T2DM, n (%)  No T2DM, n (%) | 29/44 (65.9)  322/413 (78) | 0.072 | 28/38 (73.7)  288/332 (86.7) | 0.031 | 1/6 (16.7)  34/81 (42) | 0.223 |
| *Duration of T2DM* | | | | | | |
| T2DM Duration < 5 years, n (%)  T2DM Duration 5-10 years, n (%)  T2DM Duration > 10 years, n (%) | 4/6 (75)  18/27 (66.7)  7/11 (63.6) | 0.349 | 4/6 (75)  17/22 (77.3)  7/10 (70) | 0.155 | 0 (0)  1/5 (20)  0/1 (0) | 0.443 |
| *Control of T2DM* | | | | | | |
| T2DM-Optimum control, n (%)  T2DM-No optimum control, n (%) | 28/43 (65.1)  1/1 (100) | 0.141 | 27/37 (73)  1/1 (100) | 0.073 | 1/6 (16.7)  0 | 0.223 |
| *T2DM management* | | | | | | |
| Monotherapy, n (%)  Combination therapy, n (%)  Insulin, n (%)  No medication, n (%) | 11/13 (84.6)  15/26 (57.7)  0  3/5 (60) | 0.076 | 10/11 (90.9)  15/23 (65.2)  0  3/4 (75) | 0.035 | 1/2 (50)  0/3  0  0/1 | 0.411 |
| *Hypertension (HTN)* | | | | | | |
| HTN, n (%)  No HTN, n (%) | 53/86 (61.6)  298/371 (80.3) | <0.001 | 47/67 (70.1)  269/303 (88.8) | <0.001 | 6/19 (31.6)  29/68 (42.6) | 0.384 |
| *Duration of HTN* | | | | | | |
| HTN Duration < 5 years, n (%)  HTN Duration 5-10 years, n (%)  HTN Duration > 10 years, n (%) | 16/27 (59.3)  26/41 (63.4)  11/18 (61.1) | 0.003 | 16/24 (66.7)  20/26 (76.9)  11/17 (64.7) | 0.001 | 0/3 (0)  6/15 (40)  0/1 (0) | 0.414 |
| *HTN Management* | | | | | | |
| RAAS Blockers, n (%)  CCBs, n (%)  Combined, n (%)  No Medicine, n (%) | 23/36 (63.9)  13/20 (65)  10/17 (58.8)  1/2 (50) | 0.052 | 23/33 (69.7)  13/18 (72.2)  10/15 (66.7)  1/1 (100) | 0.003 | 0/3 (0)  0/2 (0)  0/2 (0)  0/1 (0) | 0.204 |
| *Dyslipidemia* | | | | | | |
| Dyslipidemia, n (%)  No Dyslipidemia, n (%) | 18/21 (85.7)  333/436 (76.3) | 0.523 | 18/21 (85.7)  298/349 (85.3) | 0.917 | 0  35/87 (40.2) | * |
| *Ischemic heart disease (IHD)* | | | | | | |
| IHD, n (%)  No IHD, n (%) | 9/11 (81.8)  342/446 (76.7) | 0.690 | 9/11 (81.8)  307/359 (85.5) | 0.666 | 0  35/87 (40.2) | * |
| *Types of Vaccines* | | | | | | |
| Covishield, n (%)  Covaxin, n (%) | 362/370 (97.8)  69/87 (79.3) | <0.001 |  |  |  |  |
| p<0.05 considered as statistically significant, p computed by chi-square test, BMI- body mass index, T2DM- type 2 diabetes mellitus, HTN- hypertension, RAAS- Renin angiotensin aldosterone system, CCB- calcium channel blocker, IHD- ischemic heart disease, COVID-19- coronavirus disease 2019, * - No statistics is computed | | | | | | |

**Supplementary Table 2: Median (interquartile range) anti-spike antibody after the second dose of either vaccine in SARS-CoV-2 naïve cohorts**

| Characteristics | Overall, N=457 | | Covishield, N=370 | | Covaxin, N=87 | |
| --- | --- | --- | --- | --- | --- | --- |
|  | Antibody Titer,  Median (IQR)  (in AU/mL) | P | Antibody Titer,  Median (IQR)  (in AU/mL) | P | Antibody Titer,  Median (IQR)  (in AU/mL) | P |
| *Age* | | | | | | |
| Age ≤ 60 years  Age > 60 years | 113 (66.25-188)  87 (28-153) | 0.027 | 118 (79-205)  90 (31-153) | 0.033 | 52 (26.5-126)  36 (8.5-127) | 0.285 |
| *Sex* | | | | | | |
| Male  Female | 99.5 (51-166)  121 (73-196) | 0.012 | 112 (66-190)  127 (87-226) | 0.024 | 45 (19-88)  71.5 (30.5-146) | 0.187 |
| *Body-mass Index (BMI)* | | | | | | |
| BMI < 25 kg/m^2^  BMI 25-29.9 kg/m^2^  BMI ≥ 30 kg/m^2^ | 113 (56.5-188)  107 (55-173)  104 (68.5-171.75) | 0.295 | 116.5 (77-204)  113 (68.5-175)  109 (71-205.5) | 0.342 | 45 (20.75-128.5)  63 (15-140.5)  71 (30.5-108.5) | 0.702 |
| *Blood group* | | | | | | |
| A+ve  B+ve  AB+ve  O+ve  A-ve  B-ve  AB-ve  O-ve | 120.5 (70.75-325)  133.5 (67.25-266)  123 (52.5-243)  107 (58.5-199)  110 (91-292)  200 (61-326)  79 (63-79)  177 (88-400) | 0.271 | 150 (82.25-350.75)  137 (72.25-297.5)  125 (62.25-252.75)  155 (84-207)  110 (91-292)  79 (63-79)  185.5 (108.25-400) | 0.320 | 43.5 (13.75-102.75)  62 (29.25-146)  69 (4-112.5)  53.5 (18.5-125.5)  NA  NA  NA  65 (65-65) | 0.737 |
| *Comorbidities* | | | | | | |
| Any Co-morbidities  No Co-morbidities | 104 (50.5-162)  111 (66-188) | 0.373 | 113 (55-175)  118 (80-208) | 0.280 | 23 (4-52)  55 (25.75-128.5) | 0.019 |
| *Type 2 diabetes mellitus (T2DM)* | | | | | | |
| T2DM  No T2DM | 109 (62.5-179.5)  113 (33.75-195.5) | 0.121 | 114 (61.75-202.5)  115 (77-199) | 0.169 | 4.5 (4-76.25)  54 (24-126) | 0.049 |
| *Duration of T2DM* | | | | | | |
| T2DM Duration < 5 year  T2DM Duration 5-10 year  T2DM Duration > 10 year | 114 (91.75-262.75)  88 (13-161)  190 (28-364) | 0.739 | 114 (91.75-262.75)  94 (61.75-179.75)  193 (45.25-373) | 0.927 | NA  5 (4-101.5)  4 (4-4) | 0.655 |
| *Control of T2DM* | | | | | | |
| T2DM-Optimum control  T2DM-No optimum control | 113 (28-194)  213 (213-213) | 0.731 | 114 (59.5-197.5)  213 (213-213) | 0.565 | 4.5 (4-76.25)  NA | * |
| *T2DM Management* | | | | | | |
| Monotherapy  Combination therapy  Insulin  No medication | 153 (79.5-205.5)  100.5 (19-196.75)  NA  64 (28.5-276) | 0.287 | 161 (100-217)  114 (28-199)  NA  57.5 (17.25-316) | 0.225 | 27.5 (4-27.5)  4 (4-4)  NA  152 (152-152) | 0.221 |
| *Hypertension (HTN)* | | | | | | |
| HTN  No HTN | 88 (44.75-137.25)  113 (66-190) | 0.001 | 104 (58-161)  118 (80-207) | 0.001 | 51 (10-92)  54.5 (25.75-135) | 0.366 |
| *Duration of HTN* | | | | | | |
| HTN Duration < 5 years  HTN Duration 5-10 years  HTN Duration > 10 years | 90 (49-131)  85 (41-124)  113.5 (43.75-195.25) | 0.717 | 107 (61.75-132.5)  88 (52.25-179.75)  115 (53-196.5) | 0.001 | 35 (23-35)  51 (10-98)  5 (5-5) | 0.366 |
| *HTN Management* | | | | | | |
| RAAS Blockers  CCBs  Combined  No Medicine | 87 (45.25-152.25)  82.5 (38.25-195.5)  113 (61.5-124)  75.5 (20-75.5) | 0.772 | 90 (52-157)  93.5 (43.75-237.25)  114 (77-125)  131 (131-131) | 0.615 | 23 (5-23)  42 (35-43.5)  24 (4-27.5)  20 (20-20) | 0.800 |
| *Dyslipidemia* | | | | | | |
| Dyslipidemia  No Dyslipidemia | 88 (46.5-141.5)  213 (213-213) | 0.716 | 88 (46.5-141.5)  213 (213-213) | 0.862 | NA  20 (51-125) | * |
| *Ischemic heart disease (IHD)* | | | | | | |
| IHD  No IHD | 115 (76-200)  110.5 (59-183) | 0.569 | 88 (71-161)  115 (76-200) | 0.365 | NA  20 (51-125) | * |
| *Types of vaccines* | | | | | | |
| Covishield  Covaxin | 115.5 (75.75-199.25)  51 (20-125) | <0.001 |  | | | |
| p<0.05 considered as statistically significant, p computed by Mann-Whitney test or Kruskal-Wallis test, BMI- body mass index, T2DM- type 2 diabetes mellitus, HTN- hypertension, RAAS- Renin angiotensin aldosterone system, CCB- calcium channel blocker, IHD- ischemic heart disease, COVID-19- coronavirus disease 2019, * - No statistics is computed | | | | | | |

**Supplementary Table 3:** **Multiple regression to identify the independent predictors for non-responder rate (log-transformed IgG levels were used as outcome variable): -**

| Model | | Unstandardized Coefficients | | Standardized Coefficients | t | P value | 95.0% Confidence Interval for B | |
| --- | --- | --- | --- | --- | --- | --- | --- | --- |
|  |  | B | Std. Error | Beta |  |  | Lower Bound | Upper Bound |
| FINAL STEP | (Constant) | 3.231 | .185 |  | 17.45 | <0.001 | 2.868 | 3.595 |
|  | AGE | .038 | .067 | .037 | .571 | .568 | -.094 | .171 |
|  | SEX (ref=male) | .085 | .036 | .093 | 2.370 | .018 | .015 | .155 |
|  | BMI | -.001 | .025 | -.001 | -.031 | .975 | -.049 | .048 |
|  | BLOOD GROUP | .000 | .012 | .001 | .026 | .980 | -.023 | .023 |
|  | ASSOCIATED COMORBIDITY | -.133 | .052 | -.106 | -2.573 | .010 | -.235 | -.031 |
|  | IHD | .072 | .134 | .025 | .535 | .593 | -.192 | .336 |
|  | Dyslipidaemia | 1.156E-005 | .000 | -.041 | -.901 | .368 | .000 | .000 |
|  | Vaccine Type (ref=Covishield) | -.387 | .047 | -.327 | -8.244 | <0.001 | -.479 | -.295 |
|  | No Past H/O Covid19 infection  Pre-vaccination | -.450 | .056 | -.316 | -8.103 | <0.001 | -.559 | -.341 |
| a. Dependent Variable: Log (Antibody Titre), a-coefficient | | | | | | | | |

**Supplementary table 4: People who had COVID-19 after first and second dose of both vaccines among SARS-CoV-2 naïve cohorts (N = 492).**

|  | N,  (%) | Suspected | Confirmed | After first dose but before second dose | | | After second dose | | | | Median anti-spike antibody titre after first dose  (AU/mL) | Median anti-spike antibody titre after second dose  (AU/mL) | Severity  (Mild/  Moderate/  severe) | Outcome  (Recovered/ death) |
| --- | --- | --- | --- | --- | --- | --- | --- | --- | --- | --- | --- | --- | --- | --- |
|  | | | | ≤ 2-weeks | >2 – 4-weeks | > 4-weeks | ≤ 2-weeks | >2 – 4-weeks | >4 – 6-weeks | >6-weeks |  | | | |
| Total | 30,  (6.1) | 4,  (All HRCT+) | 26,  (4, RAT+;  22, RT-PCR+) | 0 | 1 | 2 | 3 | 2 | 11 | 11 | 37.5 | 99.5^@^ | Mild: 28,  Moderate: 2,  Severe: 0 | Recovered: 30,  Death: 0 |
| Covishield | 25,  (6.3) | 2 | 23 | 0 | 0 | 0 | 3 | 2 | 10 | 10 | 41.2 | 104.0 | Mild: 23,  Moderate: 2,  Severe: 0 | Recovered: 25,  Death: 0 |
| Covaxin | 5,  (5.4) | 2 | 3 | 0 | 1 | 2 | 0 | 0 | 1 | 1 | 3.8 | 42.3^$^ | Mild: 5,  Moderate: 0,  Severe: 0 | Recovered: 5,  Death: 0 |
| ^@^ 28 cohorts; ^$^ 3 cohorts; | | | | | | | | | | | | | | |
