## Supplementary figures and images for "Antibody Response after Second-dose of ChAdOx1-nCOV (Covishield™^®^) and BBV-152 (Covaxin™^®^) among Health Care Workers in India: Final Results of Cross-sectional Coronavirus Vaccine-induced Antibody Titre (COVAT) study"

### Supplemental Figure 1

**Supplementary figure 1:**

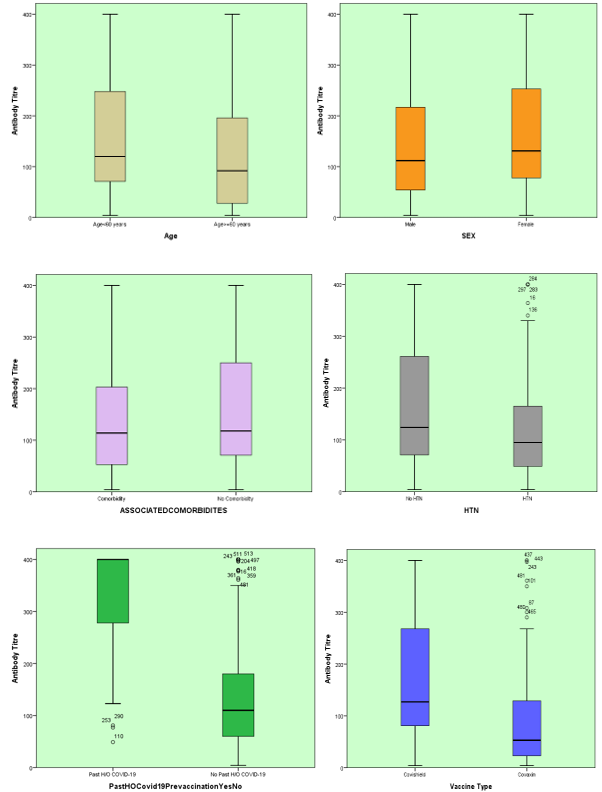
